# Loneliness and Cardiovascular Disease Risk: A Cross-National Study of Two Nationally Representative Cohorts of Older Adults in the US and South Korea

**DOI:** 10.1101/2023.10.20.23297341

**Authors:** Harold H. Lee, Ruijia Chen, Sakurako S. Okuzono, Laura D. Kubzansky

**Affiliations:** Department of Biobehavioral Health, The Pennsylvania State University, State College, PA, USA; Department of Epidemiology, Boston University School of Public Health, Boston, MA, USA; Department of Social & Behavioral Sciences, Harvard T.H. Chan School of Public Health, Boston, MA, USA

**Author notes:** <u>Correspondence and reprint requests to</u> Harold H. Lee, Department of Biobehavioral Health, College of Health and Human Development, The Pennsylvania State University, State College, Pennsylvania, 124 Biobehavioral Health Building, University Park, PA 16802, USA.

## Abstract

**BACKGROUND:** Epidemiological studies demonstrate higher loneliness is associated with increased risk of developing cardiovascular disease (CVD). However, most studies are conducted with populations in Western developed countries, whose cultures generally emphasize independence. Less clear is whether these associations are also evident in more interdependent cultures, such as those in East Asian countries. We hypothesized feeling lonely could be more stressful and exhibit stronger associations with CVD risk in a highly interdependent versus independent culture.

**METHODS:** We examined associations of loneliness with fatal and non-fatal CVD incidence in older adults from the Health and Retirement Study (HRS; n =13,073) conducted in the U.S. and from the Korean Longitudinal Study of Aging (KLoSA; n=8,311) conducted in South Korea. In both cohorts, baseline loneliness was assessed using one item from the Center for Epidemiologic Studies Depression Scale. Incident CVD was defined as reporting new-onset CVD on the biennial questionnaire or CVD death reported by proxies. Within each cohort, we estimated adjusted hazard ratios (aHR) of incident CVD according to loneliness (yes/no) over 12-14 years of follow-up, adjusting for relevant baseline covariates, including social isolation, sociodemographic factors, health conditions, and health behaviors. We further examined health behaviors as a potential pathway underlying these associations using counterfactual mediation analyses.

**RESULTS:** Controlling for all covariates, feeling lonely was associated with an increased likelihood of developing CVD in the U.S. (aHR:1.15, 95%CI: 1.04,1.27) and in South Korea (aHR: 1.16, 95%CI: 1.00, 1.34). The pooled analysis showed no heterogeneity (Q=0.009, p=0.92), and the HR for loneliness was 1.14 (95% CI: 1.05-1.23). Regarding potential mediators, several behaviors accounted for a proportion of the association: physical activity, in both countries (14.6%, p=0.03 in HRS; 1.3%, p = 0.04 in KLoSA), alcohol consumption only in KLoSA (1.1%, p < 0.001), smoking only in HRS (4.7%, p < 0.001).

**CONCLUSIONS AND RELEVANCE:** Contrary to our hypothesis, the magnitude of the loneliness-CVD relationship was similar in both countries, with 14% higher odds of developing CVD, while behavioral pathways appeared different. Loneliness may be a risk factor for CVD regardless of culture; however, different prevention strategies in clinical settings may be required.

**Clinical Perspective:** **What is New?**

- Even after controlling for social isolation, health behaviors/conditions, and sociodemographic factors, feeling lonely was associated with an increased likelihood of developing CVD among older adults in both the U.S. (15% increase) and South Korea (16% increase).
- The impact of loneliness on CVD risk did not appear to differ substantially by culture, comparing individuals from a more independent versus interdependent culture.
- The behaviors linking loneliness and CVD differed somewhat between the U.S. and South Korea, suggesting cultural factors may contribute to shaping distinct behavioral pathways through which loneliness impacts health.

**What are the clinical implications?**

- A consistent association between loneliness and CVD risk was evident in two very different cultures, suggesting loneliness may be a relevant target for CVD prevention strategies in diverse populations.
- While the associations are modest, the public health implications of loneliness-related CVD could be significant if a substantial portion of the population experiences loneliness, particularly in the aftermath of the COVID-19 pandemic.
- Assessing loneliness levels may provide healthcare professionals with greater insight into patients’ CVD risk.

## Introduction

Cardiovascular diseases (CVD) are the leading cause of death in developed countries worldwide.^1^ In 2020, approximately 19 million deaths were attributed to CVD globally, which was an increase of 18.7% from 2010.^2^ Extensive epidemiological and biomedical research on CVD has identified a number of biobehavioral risk factors, including diet, physical activity, tobacco use, harmful use of alcohol in addition to conventional risk factors, such as family history of disease, hypertension, or diabetes.^3–5^ However, these risk factors do not fully account for disease burden and more recent research indicates that various forms of psychological distress, including loneliness, may increase the risk of developing CVD.^6,7^

The recognition of loneliness as a notable public health concern grew with the appointment of a Loneliness Minister in the UK, a sentiment further emphasized by the widespread experience of extended social distancing during the COVID-19 pandemic, and subsequently reaffirmed as a priority by the U.S.. Surgeon General.^8,9^ Loneliness refers to feeling there is a discrepancy between one’s desired and actual social relationships. While related to social isolation, loneliness differs in that it arises from a *sense* of being socially isolated, which can be experienced even by those with objective evidence of social connections (e.g., being married, having frequent contact with family and friends).^10,11^ A systematic review of empirical studies examining the association between loneliness and CVD published through 2015,^12^ identified only four longitudinal studies. Except for one study with a small sample size (n<500),^13^ loneliness was consistently associated with a higher risk of developing CVD, and these associations were generally evident even after controlling for conventional risk factors and a variety of potential confounders, such as age, systolic blood pressure, body mass index, diabetes, cigarette smoking, education, income, and race.^14–16^ These findings have been corroborated by several recent well-powered longitudinal studies published in the intervening time, including 479,054 men and women from UK Biobank (age 40-69 years),^17^ 57,825 older women in the US (age 65 to 99 years),^18^ and a nationally representative sample of people aged 50 and over living in England.^19^ However, all studies were conducted in Western countries, limiting the finding’s generalizability. Research must examine the extent to which findings ascertained in these populations may (or may not) apply to individuals from other countries and cultures.^20^

The degree to which loneliness increases the risk of developing CVD may vary depending on culture and related social norms. Such differences may be particularly pronounced if cultures vary substantially in the nature of sociality. Prior research suggests Western and East Asian countries differ significantly in how individuals relate to each other. For example, people in East Asian cultures are more interdependent and more likely to define themselves in relation to others relative to individuals in Western cultures.^21–23^ Therefore, it is reasonable to hypothesize that the associations between loneliness (or other factors related to social connectedness) and CVD incidence may differ by country, and be stronger in countries with high levels of interdependence. This study aimed to examine the association between loneliness and CVD incidence among older adults in two countries characterized by these different cultures, the U.S. and South Korea representing Western and East Asian cultures respectively. Comparing advanced nations such as South Korea and the United States offers a unique opportunity to explore how cultural differences affect the link between loneliness and overall health, with less emphasis on economic factors. We hypothesized that loneliness would be associated with increased risk of incident CVD in both countries, but that loneliness would be more strongly associated with CVD incidence in South Korea than in the U.S.. We examined social isolation as a covariate as well as a potential effect modifier, given prior work suggesting it is related to loneliness but captures a different experience that can separately influence health.^24,25^ Similarly, we explored sex in these roles due to the notable loneliness-CVD link in women.^15,18^ We also examined health behaviors as potential mediators of loneliness-CVD relationships to obtain insight into how the perception of loneliness could be linked to an increased risk of CVD.

## Methods

### Participants

Data were obtained from the Health and Retirement Study (HRS) and Korean Longitudinal Study of Aging (KLoSA) obtained from harmonized data files in the Gateway to Global Aging Data, a National Institute of Health-funded public resource designed to facilitate cross-national and longitudinal studies on aging worldwide. The HRS is a panel study started in 1992 that surveys a representative sample of U.S. adults over the age of 50, and the Korean Longitudinal Study of Aging (KLoSA) is a sister study started in 2006, intentionally designed to be comparable with the HRS.^26^ Both the HRS and the KLoSA gather data on aging-related health outcomes and changes in health status over time through questionnaire assessments administered at 2-year intervals. Follow-up data including CVD mortality are available in KLoSA through 2018, and in HRS until 2014.^a^ To ensure a comparable length of follow-up and general time frame with the KLOSA data, HRS data from waves undertaken during the 2002-2014 period were included, thereby enabling a comprehensive examination of CVD mortality over an approximately 12-year period. Individuals who had already developed CVD prior to this study’s baseline (2002 in the HRS and 2006 in the KLoSA) and participants with missing data on the loneliness question in the CESD at baseline were excluded. Among participants who did not develop CVD, those who did not provide data related to CVD incidence were excluded. The final analytic sample included 13,073 American adults from the HRS and 8,311 South Korean adults from the KLoSA (see Figure 1 for sample details).

**Figure 1.**
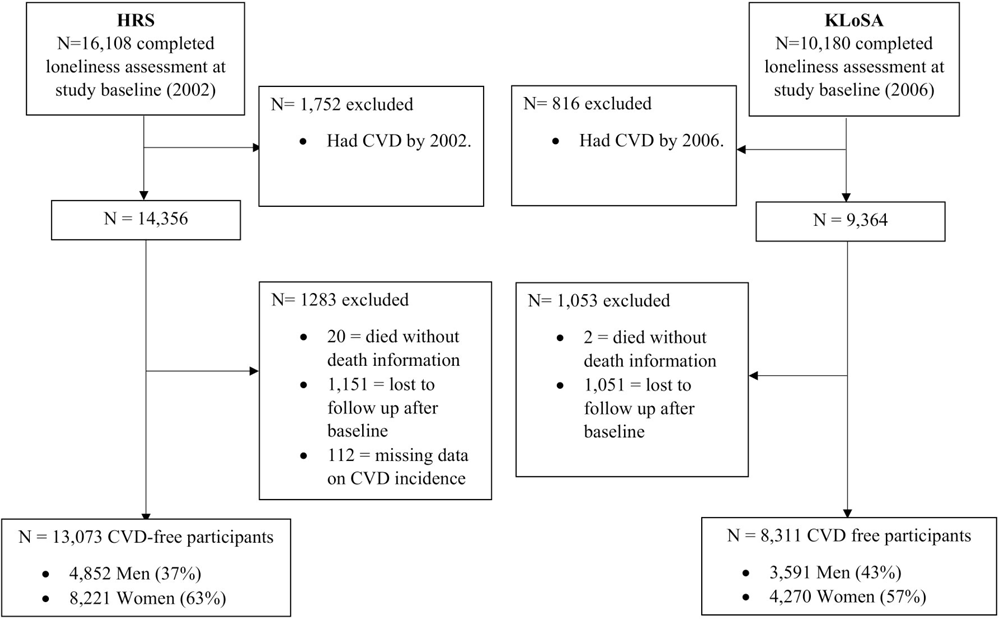
Flow diagram of subject inclusion and exclusion in HRS (2002-2014) and KLoSA (2006-2018)

### Measures

#### Loneliness

To facilitate the cross-national comparison, loneliness was operationalized using the same single question about loneliness included in the Center for Epidemiologic Studies Depression (CES-D) scale administered at our study baseline in both the HRS (2002) and KLoSA (2006). While the UCLA loneliness questionnaire was measured in a subset of HRS participants, it was obtained from only ∼15% of the HRS participants in 2002, and it was not administered in KLoSA. However, the single-item measure of loneliness from the CES-D correlates highly with other validated loneliness scales (e.g., the 20-item original UCLA scale of loneliness^27^) and has been shown to be valid.^28,29^ In HRS, participants were asked to respond to the item, ‘I felt lonely’, with a binary response option (yes/no). In KLoSA, participants were asked to respond to the item ‘I felt lonely’ with a 4-point response regarding the frequency of occurrence (0 = none or almost none; 1 = some of the time, 2 = most of the time, and 3 = All or almost all). To facilitate a direct comparison with HRS, we recategorized this as a binary variable (0 vs. 1, 2, and 3).

#### Incident cardiovascular disease

Incident CVD was characterized according to reports of new-onset CVD or CVD mortality. In both the HRS and the KLoSA, incident CVD was assessed from questions asking whether participants were diagnosed with heart attack, coronary heart disease, angina, congestive heart failure, other heart problems, or stroke and was included in all questionnaires administered at two-year intervals from 2006-2018 in KLoSA and 2002-2014 in HRS. A high correlation between self-reported CVD incidence in these types of assessments and hospital records has been well documented,^30,31^ and a previous HRS study confirmed that self-reported stroke is suitable for studying stroke and stroke risk factors.^32^ In HRS and KLoSA, cause of death data was obtained by proxies who were previously identified as close friends or relatives of the deceased when available and by vital statistics agencies in each country (i.e., National Death Index in the US and National Statistical Office in South Korea).

#### Covariates

Sociodemographic factors include age (year) and sex (male, female). Race, obtained only in HRS, was assessed with the question “What race do you consider yourself to be: White, Black or African American, American Indian, Alaska Native, Asian, Native Hawaiian, Pacific Islander, or something else?” We then recategorized this into 3 categories: Whites, Black/African Americans, and others due to the small sample size in all remaining categories. Ethnicity, also obtained only in HRS, was assessed with the question, “Do you consider yourself Hispanic or Latino?” and coded as Hispanic or non-Hispanic. In HRS and KLoSA, educational attainment was self-reported from 0 to 17 years and then recategorized into 3 categories: college graduate, high school graduate, and less than high school. Income and wealth were assessed in U.S. dollars and Korean won in HRS and KLoSA, respectively, were categorized into quartiles for harmonization. Income in HRS is the sum of the respondent’s wage, salary income, bonuses, overtime pay, commissions, tips, 2^nd^ job or military reserve earnings, and professional practice or trade income. In KLoSA, income includes the after-tax income from wage income and second job earnings. In HRS and KLoSA, total wealth is calculated as the sum of all wealth components minus all debt (KLoSA: total mortgage, total debt, lease safety deposit, safety deposits for other real estate; HRS: total mortgage, home equity line of credit balances and home equity loans, other debts, 2^nd^ home mortgage).

For CVD risk-related health behaviors and health conditions, which can be confounders or mediators in the loneliness-CVD relationship,^33,34^ we included diabetes, hypertension, body mass index (BMI; kg/m^2^), smoking, frequency of alcohol consumption, and physical activity, all assessed via self-report on the 2006 questionnaire in each sample. Prior research has validated these health condition measures,^35^ and showed substantial agreement between self-reported health conditions and medical records.^30^ Diabetes and hypertension status were determined in both HRS and KLoSA by asking participants whether a doctor had ever diagnosed them with either condition (yes/no). In HRS and KLoSA, self-reported height and weight were used to derive BMI as a continuous variable (kg/m^2^). Cigarette smoking status was assessed in both HRS and KLoSA: current, former, and latter. The frequency of alcohol consumption was measured by querying the number of days per week drinking alcohol; response options were 0-7 in HRS and 0 (none or < once a month), 1(one to several times a month), 2 (one to several times a week), 3 (most days of the week), and 4 (every day of the week) in KLoSA. In KLoSA, the frequency of exercise participation per week, regardless of intensity, was assessed. In HRS, vigorous physical activity participation (3 or more times a week; yes/no) was assessed.

Social isolation was assessed using the Berkman-Syme Social Network Index^36^ administered in HRS and KLoSA at the study baseline, using information from 4 domains: 1) marital status, 2) contact with close friends and relatives, 3) participation in group activities, and 4) participation in religious activities. Consistent with prior work,^37^ we assigned a score of 0 to those who were the least integrated and a score of 3 to those who were the most integrated within each domain, as described in more detail (Supplement. Table A). Domain scores were summed (range:0–12) to derive an overall social isolation score used as a continuous variable, whereby lower scores indicated being more socially isolated.

In KLoSA, all covariates had a low missingness (<3%). In HRS, most covariates had low missingness (<3%), except for some subcomponents of the social network index, such as religious activity participation (16%), group participation (19%), and contact with friends and family (18-23%). For all missing covariates, we conducted multiple imputations (n=5) by chained equations, which tends to be more flexible than other methods for handling missing data.^38,39^ This method also helps address problems associated with attrition.^40,41^ The imputation was performed using the MICE package in R and combined results from each imputed dataset using the MITOOL package in R.^42^

### Statistical analyses

Analyses were conducted separately in KLoSA and HRS and then compared across cohorts. While merging datasets and using the country as an effect modifier is possible, it risks oversimplification due to heterogeneity in how variables were assessed (detailed in the method section); to compute a meta-analytic effect across two cohorts, the estimates were pooled using the *metafor* package in R.^43^

We first examined the distribution of covariates by loneliness level within each dataset separately using the Pearson chi-square test for categorical variables and the t-test for continuous variables. To examine the loneliness-CVD incidence relationship, we used Cox proportional hazards models using 4 sets of models. The first model adjusted for age. The second model was further adjusted for sociodemographic factors that may confound the loneliness-CVD association, including sex, income, wealth, education, and race/ethnicity (race/ethnicity was measured in HRS, but not KLoSA). The third model additionally controlled for social isolation. Finally, the fourth model additionally included diabetes status, hypertension status, BMI, smoking status, alcohol consumption, and physical activity level, each of which can either confound or mediate the association of interest. The Cox models’ proportional hazards assumption for loneliness, which was examined using Schoenfeld residuals,^44^ was upheld in all models in both countries (*p >* 0.05).

We also conducted formal mediation analyses considering each health behavior as a potential mediator, adjusting for sociodemographic factors in the second model. To overcome the limitations of traditional mediation approaches with low statistical power,^45,46^ we employed a counterfactual mediation model using the R package *regmedint*,^47^ which was developed based on SAS macro by Valeri and Vanerweele.^48^ This model is flexible and yields higher statistical power by incorporating an exposure-mediator interaction term (e.g., loneliness × smoking).^49^ As loneliness was measured before the health behaviors were assessed, we satisfied the assumption of temporal ordering. Notably, in 2004, HRS began gathering data on light to moderate-intensity physical activity (3 or more times a week), which was used to create an intensity-weighted sum score (light=1, moderate=2, vigorous=4; range 0-49)^50–52^ for the mediation analysis.

We conducted several secondary analyses. First, we evaluated potential effect modification by social isolation. To simplify the interpretation of interactions, we dichotomized the Berkman-Syme Social Network Index as a binary variable (i.e., “isolated” vs. “not isolated”), using cut points that distinguish ∼ 15 % of the most socially isolated individuals in both countries (i.e., Berkman-Syme Social Network Index score <4). Second, we evaluated potential effect modification by sex. We tested additive interaction by calculating the relative excess risk due to interaction (RERI)^53^ and multiplicative interaction by adding a product term of to the Cox proportional hazards model adjusting for sociodemographic factors (i.e., model 2). We also calculated E-values^54^ to quantify the extent to which an unmeasured confounder would need to be associated with both social integration and mortality to explain away the observed loneliness- CVD risk association. Second, to reduce concerns about reverse causation (i.e., underlying illness could cause changes in loneliness), we conducted an analysis that excluded 1,313 HRS participants and 290 KLoSA participants who developed CVD within the first 4 years of study from baseline (resulting in n=11,891 in HRS and n=8,091 in KLoSA). Third, instead of imputing data, we conducted complete case analyses excluding 5,402 HRS participants (41%) and 266 KLoSA participants (3%) missing any component of the covariate measures (resulting in n = 7,802 in HRS and 8,045 in KLoSA).

All analyses were conducted using R™ (Team, 2016). To account for design effects in each cohort, appropriate survey weights were applied using the *svy* command. Additive interaction analyses were performed using *the interactionR* package ^55^, with the delta method for computing the 95% confidence interval.^56^ Because survey weights cannot be applied in *regmedint* or *interactionR*, causal mediation and additive interaction analyses were performed without survey weights.

## Results

Compared to South Koreans in KLoSA, Americans in the HRS were older at baseline (mean age 66 ± 9 years vs. 57 ± 10 years) and were better educated (e.g., percentage of college graduate = 24% vs. 11%). In general, South Korean adults had a lower prevalence of adverse health conditions and better health behaviors than American adults. For example, comparing adults in South Korea and the U.S. at baseline, the prevalence of diabetes was 10% versus 14%; the prevalence of hypertension was 22% versus 49%; the prevalence of never smoking was 69% versus 43%; respectively.

At baseline, 16% of the adults in the U.S. sample and 24% of the adults in the South Korean sample felt lonely. During the 12-year follow-up, 25% of HRS and 11% of KLoSA participants developed CVD. The average Social Network Index score was 6.1 ± 2.6 in HRS and 6.3 ± 2.6 in KLoSA. In both countries, those who were lonely also had lower Social Network Index scores, were more likely to be women, to be less educated, and to have a higher prevalence of diabetes and hypertension (Table 1).

**Table 1.** Sociodemographic Characteristics of a Nationally Representative Sample of U.S. adults from the Health and Retirement Study (n=13,073; and South Korean adults from the Korean Longitudinal Study of Aging (n=8,311; 2006).^a^.

|  | **HRS**<br>**(United States)** |  | **KLoSA**<br>**(South Korea)** |  |
| --- | --- | --- | --- | --- |
|  | **Not lonely**<br>**(n=11,054; 84%)** | **Lonely**<br>**(n=2,150; 16%)** | **Not lonely**<br>**(n=6,113; 76%)** | **Lonely**<br>**(2,198; 24%)** |
| **Demographics** |  |  |  |  |
| Age, mean (standard deviation), years | 66.3 (9.1) | 68.4 (10.1) | 57.2 (9.9) | 61.7 (11.3) |
| Women, % | 6,720 (58%) | 1,525 (69%) | 3,259 (50%) | 1,461 (63%) |
| Race |  |  |  |  |
| Whites, % | 9,241 (89%) | 1,646 (83%) |  | n/a |
| Black/African American, % | 1,372 (8%) | 392 (12%) |  | n/a |
| Other, % | 343 (3%) | 110 (5%) |  | n/a |
| Hispanic, % | 763 (5%) | 274 (11%) |  | n/a |
| Education |  |  |  |  |
| College graduate, % | 2,471 (29%) | 245 (16%) | 686 (13%) | 94 (5%) |
| High school graduate, % | 6,189 (57%) | 1,092 (56%) | 2,022 (38%) | 387 (22%) |
| Less than high school, % | 2,295 (14%) | 813 (28%) | 3,405 (49%) | 1,717 (72%) |
| **Health Conditions and Behaviors** |  |  |  |  |
| Diabetes, % | 1,572 (14%) | 406 (18%) | 598 (9%) | 321 (14%) |
| Hypertension, % | 5,377 (48%) | 1,162 (54%) | 1,428 (21%) | 692 (29%) |
| Body Mass Index, average (SD), kg/m<sup>2</sup> | 27.4 (5.2) | 27.5 (5.9) | 23.3 (2.6) | 23.1 (3.0) |
| Smoking, % |  |  |  |  |
| Current smoker | 1,446 (13%) | 399 (19%) | 1224 (23%) | 410 (21%) |
| Past smoker | 4,702 (44%) | 888 (39%) | 586 (9%) | 169 (8%) |
| Never smoked | 4,719 (43%) | 845 (43%) | 4302 (68%) | 1619 (72%) |
| Social Network Index<sup>b</sup>, average (SD) | 6.3 (2.6) | 4.9 (2.6) | 6.6 (2.4) | 5.3 (2.7) |

<sup>a</sup> Raw values (i.e., without imputation or survey weights) were used for the values not in parentheses. For % values in parentheses as well as continuous variables (i.e., age and body mass index), survey weights were applied to imputed data.
<sup>b</sup> Berkman-Syme Social Network Index is a composite score calculated using 1) marital status, 2) contact with friends/family, 3) group attendance, and 4) religious attendance. The score ranges from 0 to 12 with lower score indicating social isolation.

### Associations of Loneliness and CVD

Loneliness was associated with a higher risk of developing CVD in both HRS and KLoSA. While the effect size appears to be larger in HRS compared to KLoSA when only adjusting for age (1.37 in HRS vs. 1.18 in KLoSA), the effect sizes were remarkably similar across models 2 through 4 (Table 2). In the fully adjusted model, feeling lonely was associated with a 16% and 18% higher likelihood of developing CVD in KLoSA and HRS, respectively. According to the pooled analysis, there was no indication of heterogeneity (Q=0.03, *p* =0.85). From the meta-analysis of HRS and KLoSA, the pooled estimated fully adjusted HR for loneliness was 1.16, with a 95% CI from 1.07 to 1.24.

**Table 2.** Hazard Ratios for Associations of Loneliness (2002 in HRS and 2006 in KLoSA) with Major Cardiovascular Disease Incidence (2004-2014 S; 2008-2018 in KLoSA) of a Nationally Representative Sample of U.S. adults from the Health and Retirement Study (n=13,073) and South an adults from the Korean Longitudinal Study of Aging (n=8,311)

|  |  |  | Model 1<sup>b</sup> |  | Model 2<sup>c</sup> |  | Model 3<sup>d</sup> |  | Model 4<sup>e</sup> |  |
| --- | --- | --- | --- | --- | --- | --- | --- | --- | --- | --- |
|  | Person-years | Cases | HR | 95%CI | HR | 95%CI | HR | 95%CI | HR | 95%CI |
| **HRS (United States)** |  |  |  |  |  |  |  |  |  |  |
| Lonely | 16,927 | 674 | 1.31 | 1.17, 1.47 | 1.21 | 1.09, 1.35 | 1.18 | 1.07, 1.30 | 1.15 | 1.04, 1.27 |
| Not lonely | 98,909 | 2658 | 1.00 | Ref | 1.00 | Ref | 1.00 | Ref | 1.00 | Ref |
| **KLoSA (South Korea)** |  |  |  |  |  |  |  |  |  |  |
| Lonely | 21,102 | 365 | 1.18 | 1.03, 1.35 | 1.22 | 1.06, 1.40 | 1.19 | 1.03, 1.39 | 1.16 | 1.00, 1.34 |
| Not lonely | 60,507 | 778 | 1.00 | Ref | 1.00 | Ref | 1.00 | Ref | 1.00 | Ref |

<sup>a</sup> Survey weights are applied
<sup>b</sup> Adjusted for age
<sup>c</sup> Adjusted for Model 1's covariate and sex, income, wealth, education, race, and ethnicity.
<sup>d</sup> Adjusted for Model 2's covariates and social isolation.
<sup>e</sup> Adjusted for Model 2's covariates and diabetes, hypertension, body mass index, smoking, alcohol consumption, and physical activity level.

### Role of Social Isolation in Loneliness-CVD Associations

We did not find substantial evidence of a multiplicative interaction between loneliness and social isolation in either HRS (p=0.09) or KLoSA (p=0.43). Similarly, we did not find evidence of an interaction on the additive scale in either HRS (RERI=-0.06, 95% CI =-0.30 ∼ 0.17) or KLoSA (RERI: 0.04, 95% CI = -0.21 ∼ 0.3). Analyses stratified by social isolation reveal that the estimates consistently point towards a similar direction for both socially isolated and non-socially isolated individuals. However, the findings are somewhat more robust among those who were not socially isolated. (Supplement Table B).

### Role of Sex in Loneliness-CVD Associations

We did not find substantial evidence of a multiplicative interaction between loneliness and sex in either HRS (p=0.30) or KLoSA (p=0.56). Similarly, we did not find evidence of an interaction on the additive scale in either HRS (RERI=0.08, 95% CI=-0.13 ∼ 0.29) or KLoSA (RERI: -0.15, 95% CI=-0.35 ∼ 0.06). Analyses stratified by sex reveal that estimates remained largely consistent between men and women in the HRS dataset, with slightly higher estimates among men compared to women in the KLoSA dataset (Supplement Table C).

### Mediation analyses

Table 3 displays the mediation results for all hypothesized mediators, comparing people who did versus did not report feeling lonely. In summary, among the health behaviors, we found some evidence for mediation between loneliness and CVD incidence through increased physical activity particularly in HRS. While there was some evidence of mediation in KLoSA, the proportion mediated was substantially smaller (14.9%, p < 0.001 in HRS; 1.3%, p = 0.04 in KLoSA). A small proportion of the effect was also mediated through alcohol consumption in KLoSA only (1.1%, p < 0.001) and through current smoking status in HRS only (4.8%, p = 0.04).

**Table 3.** Mediation Analyses between Loneliness (2002 in HRS and 2006 in KLoSA) and Risk of CVD (2004-2014 in HRS; 2008-2018 in KLoSA) of a Nationally Representative Sample of U.S. adults from the Health and Retirement Study (n=13,073) and South Korean adults from the Korean Longitudinal Study of Aging (n=8,311)^a^.

|  | HRS<br>(United States) |  | KLoSA<br>(South Korea) |  |
| --- | --- | --- | --- | --- |
|  | Proportion mediated | p-value of PNIE | Proportion mediated | p-value of PNIE |
| Smoking | 4.7 % | <0.001 | 2.3 % | 0.92 |
| Physical activity | 14.6% | 0.03 | 1.3 % | 0.04 |
| Alcohol | 4.3% | 0.06 | 1.1% | <0.001 |

PNIE = Pure natural indirect effect expresses how much risk of developing CVD would change on average if the loneliness level was controlled (i.e., fixed) at “not lonely” but the mediator was changed from the level occurring among individuals who reported they were “not lonely” to the level occurring among individuals who reported being “lonely”.
<sup>a</sup> To ensure temporality between exposure and mediators, we used the 3 mediators (“smoker”, “physical activity”, “alcohol”) assessed at first wave of follow up after the baseline, which is approximately 2 years later.

### Sensitivity analyses

The associations between loneliness and CVD incidence in both countries were moderately robust to unmeasured confounders. For example, the *E*-value in the full model comparing lonely vs. non-lonely groups in HRS was 1.42 and 1.59 in KLoSA,^57^ suggesting that an unmeasured confounder would need to have a minimum HR of 1.42 in HRS and or 1.59 in KLoSA (with both loneliness and CVD risk) to explain away the observed association, above and beyond all of the covariates already considered.^54^ The results remained consistent when participants who developed CVD within the first 4 years (to reduce concerns about potential reverse causality) were excluded (Supplemental Table D) and when missing observations were not imputed (Supplemental Table E).

## Discussion

In this cross-national study, longitudinal associations between loneliness and CVD incidence were examined among older adults in two ongoing cohort studies, one in the U.S. and one in South Korea, each with ∼12 years of follow-up. Contrary to the hypothesis that the association would be stronger in South Koreans due to their collectivist culture, feeling lonely was associated with an increased likelihood of developing CVD in both countries, with nearly the same magnitude of 14% higher risk of CVD, controlling for social isolation, sociodemographic factors, health condition, and health behaviors. However, the behaviors linking loneliness and CVD differed somewhat between the U.S. and South Korea, suggesting cultural factors may contribute to shaping distinct behavioral pathways through which loneliness impacts health.

The sizes of the observed effects in both HRS and KLoSA for loneliness in relation to CVD are comparable with those found in prior studies. For example, when evaluating the association of loneliness with incident CVD, a sister study in England, the English Longitudinal Study of Aging, reported a 1.27 adjusted HR over 5 years, ^19^ and a systematic review including studies through 2015 revealed adjusted HRs ranging from 1.31 to 1.53^14–16^ both greater than the adjusted HR observed in the present study. It is worth noting that not all previous studies have factored in the influence of social isolation. Among those that accounted for social isolation, a trend towards smaller effect sizes emerged. For instance, a prior study within the Women’s Health Initiative, which examined the relationship between 3-item UCLA loneliness and CVD incidence in older women in the United States across a 7-year follow-up period controlling for social isolation alongside numerous covariates, reported an adjusted HR of 1.05 (95% CI=1.01- 1.09).^18^ Similarly, a previous study within the UK Biobank, exploring the association between 2- item measure of loneliness perception and CVD incidence over 8 years considering social isolation and an array of covariates, reported an adjusted HR of 1.06 (95% CI=0.96-1.17).^17^ The substantially reduced effect sizes likely indicate interconnectedness between loneliness and social isolation, with the small independent effect implies loneliness exerting a minor independent influence on cardiovascular physiology. Our analyses, combined with previous findings in Western countries,^6,7,17^ suggest that social isolation does not alter the relationship between loneliness and CVD incidence. Taken together, when controlling for a wide range of covariates, the effect of loneliness on incident CVD risk may be moderate and becomes even smaller, albeit still evident, when accounting for social isolation.

Our findings suggest potential health-damaging effects of feeling lonely (beyond those on mental health) may not be unique to Western cultures, and in fact may generalize to other cultures. In fact, the size of the association of loneliness with incident CVD risk was remarkably similar between East Asians and Westerners in our study. Moreover, the links between loneliness and CVD risk were not modified by social isolation or sex. Theses could suggest broad systemic effects that can occur via numerous pathways, which may differ across cultures. Interestingly, emerging research examining the negative impact of weak social connections on survival in wild mammals shows associations of similar magnitude to what we and others have observed in our study with humans (i.e., odds ratios ranging from 1.23 to 1.72).^58–60^ Taken together, findings in animals and humans could further suggest there may be common mechanisms linking social connection and survival, involving hormonal, vascular, immune, and behavioral responses triggered by perceived social isolation and loneliness,^33,61–63^ which are evolutionarily conserved.

In the current study, cultural differences were primarily evident in relation to the behavioral pathways by which loneliness appears to affect health. For example, alcohol consumption emerged as a mediator between loneliness-CVD incidence only in South Korea. This observation corresponds with the prevailing cultural norms in South Korea, where the consumption of alcoholic beverages enjoys broad social acceptance, extending even to public venues such as parks.^64^ Consequently, individuals experiencing loneliness within the South Korean context may exhibit diminished cultural or legal inhibitions against heavy drinking. Notably, smoking was a mediator between loneliness-CVD incidence only in the US, in which smoking has become less socially acceptable in recent decades.^65^ It is plausible that in the US context, lonely individuals experienced relatively weaker social pressure or had less motivation to conform to prevailing cultural attitudes related to smoking behavior. These findings highlight the importance of considering different mechanistic pathways in different cultures and incorporating such considerations in interventions development.

One key public health implication of these findings is that population-level interventions may consider targeting loneliness as an upstream factor that could improve cardiac health in the population. While the magnitude of the association of loneliness with cardiovascular health is smaller than some conventional CVD risk factors, most cases of disease arise in low-to-moderate risk populations rather than the smaller number of people at high risk (i.e., “prevention paradox” ^66^). Therefore, interventions that can reduce the levels of loneliness may result in a significant reduction in CVD incidence at the population level. Such findings are particularly notable given the nearly universal impact of social distancing occurring with the COVID-19 pandemic. It is not yet clear the extent to which individuals will remain feeling somewhat distant versus return to pre-pandemic levels of connectedness. Thus, population-level interventions mitigating social isolation and loneliness, or increasing emotional well-being and social integration, may be particularly relevant now. Another important question to ask other than “Should we target this risk factor?” is “Can we change it?” Two recent meta-analyses have shown that loneliness can indeed be reduced by interventions, such as cognitive behavioral therapies, social support interventions, and social and emotional skills training.^67,68^ However, it is unclear whether the decrease in loneliness will in turn lead to reductions in CVD incidence. Nonetheless, it is worth targeting an upstream factor given prior work has demonstrated how hard it is to change conventional behavioral risk factors. That is, behavioral intervention studies in the past decades have shown that some of the behavioral components of the American Heart Association Life’s Essential 8, such as diet and physical activity, are difficult to change and even more difficult to maintain the ideal level.^69,70^ Future research may investigate whether reductions in loneliness result in improved cardiovascular health.

Our study has some limitations. First, the assessment of loneliness relied on a single-item measure, although it has been validated and shown to be correlated with other loneliness scales. Second, the study design was observational, limiting our ability to establish causal relationships between loneliness and CVD. While we took measures to reduce concerns about reverse causation, including removing people who developed CVD at baseline and sensitivity analysis removing participants who developed CVD in the first 4 years, unmeasured confounding remains possible. That said, the loneliness-CVD link was still evident even after controlling for a wide array of potential confounders and mediators with moderately robust E-value. The current study also has several notable strengths. First, this is one of the few (or only) studies to compare the loneliness-CVD association in Western and non-Western populations using harmonized studies. The harmonization of data from two nations, the United States and South Korea, provides a unique opportunity for cross-cultural comparison. Further, by utilizing nationally representative data, our findings can be generalized to older adult adults in these two countries.

In conclusion, this study demonstrated that loneliness is associated with an increased risk of developing CVD in both the United States and South Korea, suggesting this association is evident even across different cultures. From a mechanistic perspective, that associations were evident even when different behavioral pathways appeared to be operating is intriguing, suggesting that loneliness may exert a systemic influence on cardiovascular health either through multiple behavioral pathways or even independent of behavior. Nonetheless, given the divergence in behavioral pathways across cultural contexts, interventions for reducing loneliness will likely need to be culturally tailored for optimal effectiveness. Future research should explore the feasibility of reducing loneliness on a population level and whether such interventions can effectively improve cardiovascular health.

## Data Availability

All of the data used for this paper is publicly available at "Gateway to Global Aging" website (here.https://g2aging.org/downloads). All analyses were ran using R, which will be available here (https://github.com/HaroldLee19/lone_cvd_us_korea.git) after publication.

https://github.com/HaroldLee19/lone_cvd_us_korea.git

## Footnotes

a Harmonized HRS End of Life Documentation (March 2019) collected data until 2014.

## Notes

### Competing Interest Statement

The authors have declared no competing interest.

### Clinical Trial

This was not a clinical trial

### Funding Statement

National Institute on Aging R24AG048024; R01AG030153).

### Author Declarations

This study is a secondary data analysis using publicly available datasets. The current work does not involve any identifiable data or HIPAA considerations.

